# Effect of emergency declaration for the COVID-19 outbreak in Tokyo, Japan in the first two weeks

**DOI:** 10.1101/2020.04.16.20067447

**Authors:** Junko Kurita, Tamie Sugawara, Yasushi Ohkusa

**Author notes:** Corresponding author: Junko Kurita.

## Abstract

**Background:** Japan’s Prime Minister Abe declared an emergency to control the COVID-19 outbreak on April 7, 2020. He asked almost half of the population of Japan to reduce their personal contacts by 70–80%.

**Object:** This study estimates the effectiveness of that emergency declaration. Method: We applied a simple susceptible–infected–recovery model to data of patients with symptoms in Tokyo, Japan for January 14 – April 21 as of April 22. We estimate the reproduction number in four periods: R_0_ before voluntary event cancellation and school closure (VECSC) which was introduced since February 27 to March 19, R_v_ during the VECSC, R_a_ after VECSC, and R_e_ after the emergency declaration.

**Results:** Results suggest that the value of R_0_ was estimated as 1.267; its range was [1.214, 1.341]. However, R_v_ was estimated as 2.360 [1.844, 2.623]. R_a_ was estimated as 2.307 [2.035, 2.794] and R_e_ was 0.462 [0.347, 0.514].

**Discussion and Concussion:** One must be reminded that these results reflect only those at two weeks after the emergency declaration. The reproduction number probably changed thereafter continuously.

## Introduction

To control the COVID-19 outbreak in Japan, the Prime Minister Abe declared an emergency on April 7 and applied countermeasures starting from April 8 [1]. The declared measures required residents of seven prefectures, collectively accounting for 44.5% of Japan’s population, to restrict their trips outside the home voluntarily. Voluntary compliance means that the declared measures were not enforced as law. People were not arrested even if they do not comply with this government declaration. At the same time, the policy required the reduction of contact by 70–80%, based on the author’s earlier research [2]. However, the share of the population cooperating with government requirements without law enforcement has remained uncertain. Therefore, this policy’s effects must be evaluated as nearly and in as real-time a manner as possible. If the reproduction number after the emergency declaration (R_e_) is less than one, then the government would be able to rescind the declaration earlier. If greater than one, and especially of greater than two, then a more powerful policy, including law enforcement, might be necessary. Therefore, we undertook preliminary evaluation of the effect as of April 14, at one week after the declaration was issued.

## Method

The COVID-19 outbreak in Japan, which occurred from January 14, and its related countermeasures, are divisible into four periods. The national government required sports and entertainment events to be canceled in Japan for two weeks from February 26 through March 19 according to a government advisory. At the same time, it was advised that small business and private meetings be cancelled voluntarily. Moreover, from March 3, almost all schools were closed to control the spread of the infection until spring vacation began in the middle of March. The policy was designated as voluntary event cancellation and school closure(VECSC). Consequently, the COVID-19 outbreak can be divided into four distinct periods: before VCSEC through February 26, VCSEC through March 19, and after VCSEC through April 7, and the after emergency was declared. We defined the reproduction numbers for the respective periods as R_0_, R_v_, R_a_, and R_e_.

Onset dates were sometimes unreported. Therefore, we estimated the onset date. To do so, we inferred a distribution of the length from onset to reporting based on patients for whom onset dates were available. Then we applied this empirical distribution to patients for whom onset dates were not available. Letting *f*(*k*) represent this empirical distribution and letting *Nt* denote the number of patients for whom onset dates were not available published at date *t*, then the number of patients for whom the onset date was *t*-1. The number of patients for whom onset dates were not available was estimated as *f*(1)*Nt*. Similarly, the number of patients with onset date *t*-2 and whose onset dates were not available was estimated as *f*(2)*Nt*. Therefore, the total number of patients whose onset date were not available, given an onset date of *s*, was estimated as Σ_*k*=1_*f*(*k*)*Ns*+*k* for a long time passed from *s*.

Moreover, the reporting delay for published data from MHLW might be considerable. In other words, if *s*+*k* was larger than that in the current period *t*, then *s*+*k* represents the future for period *t*: thus *Ns*+*k* was not observable. Such a reporting delay causes an under-estimation bias in the number of patients. Therefore, we must adjust it as Σ_*k*=1_^*t-s*^*f*(*k*)*Ns*+*k* /Σ_k=1_^*t-s*^*f*(*k*). Similarly, patients for whom the onset dates were available are expected to be affected by the reporting delay. Thus we have *Ms*|*t* /Σ_k=1_^*t-s*^*f*(*k*), where *Ms*|*t* represents the reported number of patients for whom onset dates were within period *s* extending until the current period *t*.

We applied a simple susceptible–infected–recovery (SIR) model [3,4] to the epidemic curve for Japan, with 120 million populations. We assume an incubation period calculated according to the empirical distribution.

Symptomatic and asymptomatic states continued for one week and then moved to a recovery state with probability of one. We are not concerned about outcomes or necessary medical resources in this model. Therefore, the state of death or hospitalization was not incorporated into the model. Asymptomatic cases cannot be observable unless complete laboratory-based surveillance is performed. One exceptional study found the asymptomatic cases to be 3/23 among elderly people [5]. For these analyses, the powers of infectivity among severe patients and mild patients were equal. Moreover, we assumed that asymptomatic cases have the same power of infectivity as symptomatic cases have [5]. The distribution of infectiousness in symptomatic and asymptomatic cases was assumed to be 30% on the onset day, 20% the following day, and 10% for the next five days [5].

We sought R_0_, R_v,_ R_a_, and R_e_.to fit the data to minimize the sum of absolute values of discrepancies among the bootstrapped epidemic curve and the fitted values. The estimated distribution of three reproduction numbers was calculated using 10,000 iterations of bootstrapping from the empirical distribution of the data obtained for symptomatic patients.

We estimated the results sequentially as follows: We first estimated R_0_ as the best fit to bootstrapped data in the pre-VECSC period. Then based on the obtained R_0_ and course of the outbreak in the pre-VECSC period, we estimated R_v_ as the best fit to bootstrapped data for the VECSC period. Similarly, based on the obtained R_0_ and R_v_, we estimated R_a_ as the best fit to bootstrapped data in the post-VECSC period. Finally, based on the obtained R_0_, R_v_, and R _a_, we estimated R_e_ as the best fit to a bootstrapped data after the emergency was declared. For each step, reproduction numbers were grid-searched in the interval of (0, 10) by 0.001 resolution.

### Data source

Data used were the numbers of symptomatic patients in Tokyo, Japan during January 14 – April 22 reported by Tokyo Metropolitan Government (TMG) [6] as of April 23. During this period, 6084 cases with onset dates were reported. We excluded imported cases and those who were presumed to have been infected passengers and crew from the Diamond Princess: they were presumed not to be community-acquired cases in Japan. Of those, onset dates were available for 3734 cases; onset dates were not reported for other cases.

### Ethical consideration

All information used for this study has been published. There is therefore no ethical issue related to this study.

## Results

Figure 1 depicts the empirical distribution among incubation periods of 91 cases for which exposure dates and onset dates were reported by MHLW. Its mode was six days. Its average was 6.6 days. Figure 2 depicts the empirical distribution of duration from onset to report. We adjusted the reporting delay based on this distribution.

**Figure 1:**
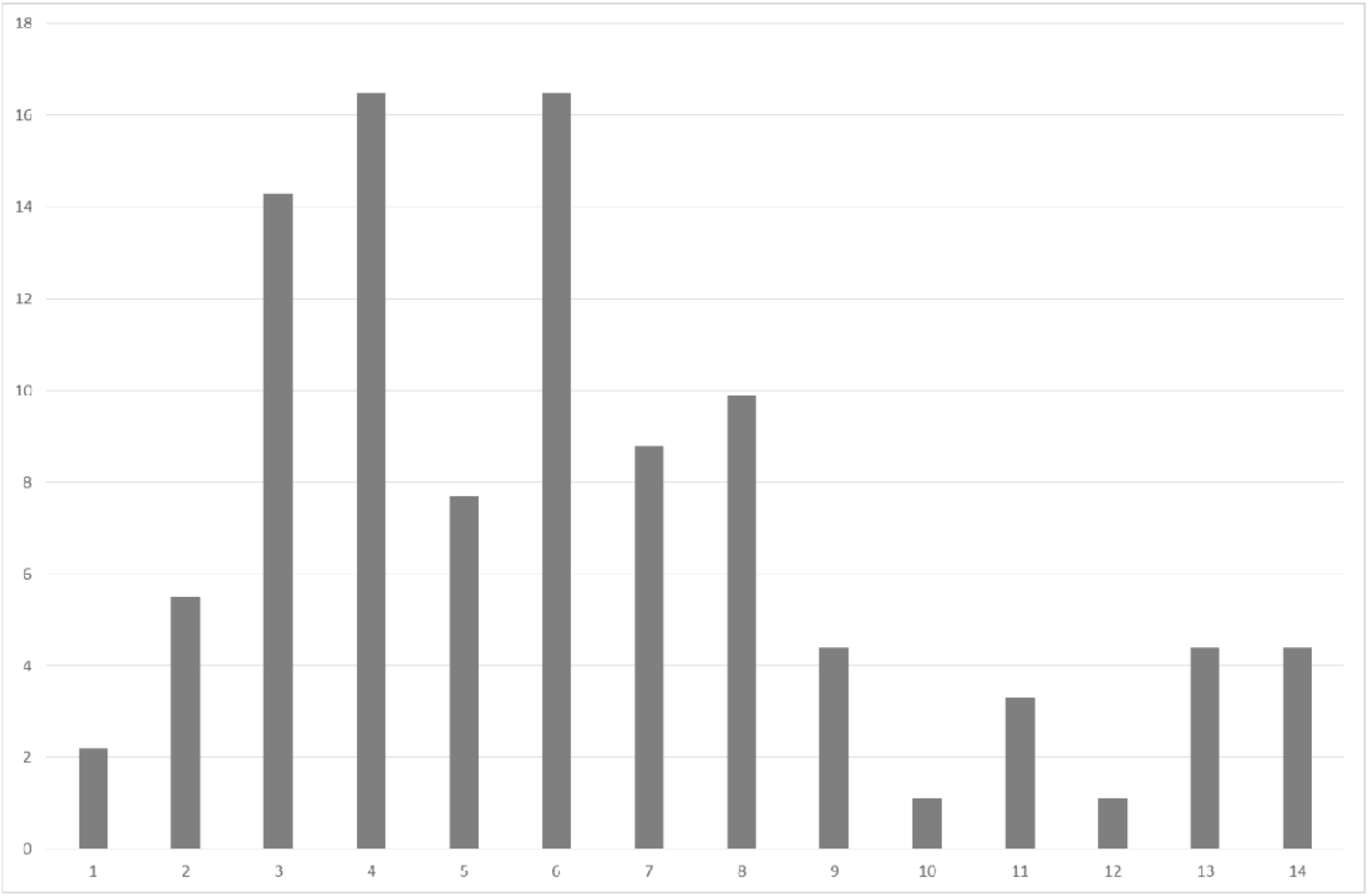
Empirical distribution of incubation period published by Ministry of Labour, Health and Welfare, Japan (%) Notes: Bars indicates the number of patients by incubation period among 59 cases whose exposed date and onset date were published by Ministry of Labour, Health and Welfare, Japan.

**Figure 2:**
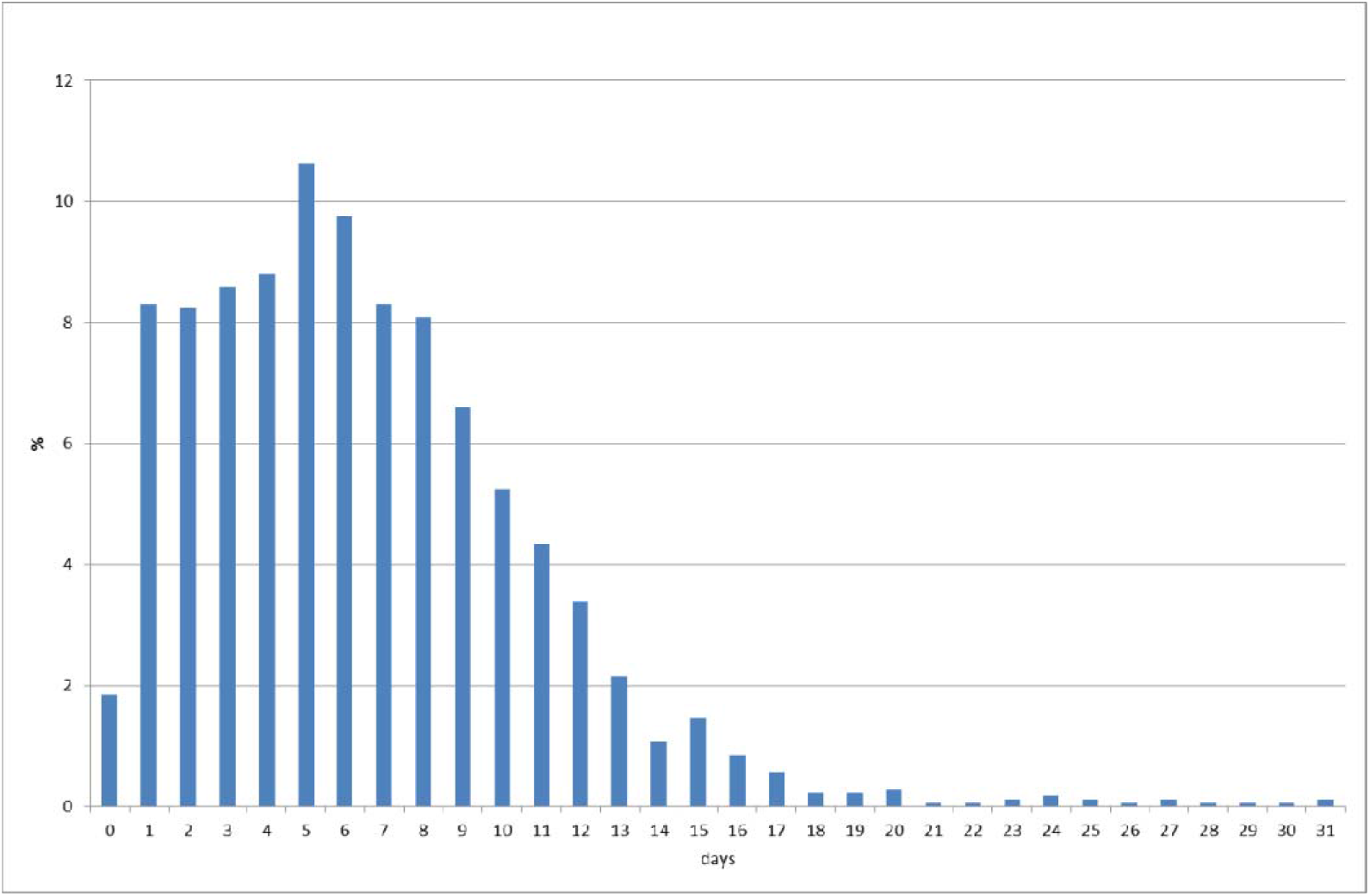
Empirical distribution of duration from onset to report by Ministry of Labour, Health and Welfare, Japan (%) Note: Bars indicates probability of duration from onset to report base on 657 patients whose onset date was available in Japan. Data source was Ministry of Labour, Health and Welfare, Japan.

The value of R_0_ before VECSC was introduced was estimated as 1.267; its range was [1.214, 1.341]. However, R_v_ during the VECSC period when measures were introduced was estimated as 2.360 [1.844, 2.623]. After VECSC, R_a_ was estimated as 2.307 [2.035, 2.794]. After the emergency declaration, R_e_ decreased dramatically to be 0.462 [0.347, 0.514]. The null hypothesis that R_v_ and R_a_ were equal was not rejected, however, all other pairs were equal was rejected at 5% significant level.

## Discussion

We applied a simple SIR model to all cases, including asymptomatic cases that had not been incorporated into the model to date. An earlier study [7–10] estimated R_0_ for COVID-19 as 2.24–3.58 in Wuhan. Our R_0_ obtained before VECSC was smaller than the previous studies, however, reproduction numbers in VECSC period and after VECSC were almost comparable. However, an earlier study [11] estimated R_0_ in Japan as 0.6 as of the end of February, which was the period mainly before the VECSC was implemented. Such a low number might mislead policy efforts for countermeasures in Japan as it adheres to contact tracing efforts to detect clusters. Outbreaks contained themselves. Emergency declarations would be unnecessary if 0.6 is correct.

Our previous study estimated R_e_ in the first a week after the emergency declaration at nationwide was 0.083 [0.067, 0.105]. However, the present study in Tokyo in the two weeks after declaration was 0.758 [0.631, 0.879] Even it was lower than one, it increased to be ten time higher from a week in nationwide. It might suggest the effect of the emergency declaration has been gradually disappearing or in Tokyo, it was higher than in nationwide.

Moreover, reproduction number in the VECSC period and after VECSC were almost the same in Tokyo, though in nationwide, reproduction number in the VECSC period (1.028 [0.965, 1.191]) was much smaller than after VECSC (3.237 [3.005, 3.441]). This might means that VECSC was ineffective in Tokyo but effective other than Tokyo. In this sense, the effect of the emergency declaration might weak in Tokyo comparison with other area.

One must be reminded that these results reflect only those at two weeks after the emergency declaration. The reproduction number probably changed thereafter continuously. Therefore, we have to monitor it carefully in the next two weeks when emergency declared and after disappeared the declaration.

## Conclusion

Results demonstrated that the reproduction number declined in the two weeks after the emergency declaration to be less than one, however, it might increase since the first week. The reproduction number must be monitored carefully and continuously. We hope that the present study contributes to government decision-making about cessation of the emergency declaration.

This study was based on the authors’ opinions. The analyses and results do not reflect any stance or policy of our affiliations.

## Data Availability

Japan Ministry of Health, Labour and Welfare. Press Releases. (in Japanese)

https://www.mhlw.go.jp/stf/newpage_10723.html

## Acknowledgments

We acknowledge the great efforts of all staff at public health centers, medical institutions, and other facilities who are fighting the spread and destruction associated with COVID-19.

